# A hemagglutination-based, semi-quantitative test for point-of-care determination of SARS-CoV-2 antibody levels

**DOI:** 10.1101/2021.05.01.21256452

**Authors:** Robert L. Kruse, Yuting Huang, Alyssa Lee, Xianming Zhu, Ruchee Shrestha, Oliver Laeyendecker, Kirsten Littlefield, Andy Pekosz, Evan M. Bloch, Aaron A.R. Tobian, Zack Z. Wang

## Abstract

Serologic, point-of-care tests to detect antibodies against SARS-CoV-2 are an important tool in the COVID-19 pandemic. The majority of current point-of-care antibody tests developed for SARS-CoV-2 rely on lateral flow assays, but these do not offer quantitative information. To address this, we developed a new method of COVID-19 antibody testing employing hemagglutination tested on a dry card, similar to that which is already available for rapid typing of ABO blood groups. A fusion protein linking red blood cells (RBCs) to the receptor-binding domain (RBD) of SARS-CoV-2 spike protein was placed on the card. 200 COVID-19 patient and 200 control plasma samples were reconstituted with O-negative RBCs to form whole blood and added to the dried protein, followed by a stirring step and a tilting step, 3-minute incubation, and a second tilting step. The sensitivity for the hemagglutination test, Euroimmun IgG ELISA test and RBD-based CoronaChek lateral flow assay was 87.0%, 86.5%, and 84.5%, respectively, using samples obtained from recovered COVID-19 individuals. Testing pre-pandemic samples, the hemagglutination test had a specificity of 95.5%, compared to 97.3% and 98.9% for the ELISA and CoronaChek, respectively. A distribution of agglutination strengths was observed in COVID-19 convalescent plasma samples, with the highest agglutination score (4) exhibiting significantly higher neutralizing antibody titers than weak positives (2) (p<0.0001). Strong agglutinations were observed within 1 minute of testing, and this shorter assay time also increased specificity to 98.5%. In conclusion, we developed a novel rapid, point-of-care RBC agglutination test for the detection of SARS-CoV-2 antibodies that can yield semi-quantitative information on neutralizing antibody titer in patients. The five-minute test may find use in determination of serostatus prior to vaccination, post-vaccination surveillance and travel screening.

## Introduction

The COVID-19 pandemic has impacted nearly all facets of health and society. The scale and speed with which SARS-CoV-2 infection spread, introduced a myriad of challenges. Early on, testing was identified as being critical to contend with the global health crisis. Testing is needed both for the diagnosis of those who are actively infected, but also for monitoring of those who have recovered. The latter is increasingly important for immune surveillance, which in turn has a range of applications spanning ascertainment of vaccination uptake to travel. This has led to a surge in the development and marketing of SARS-CoV-2 serology assays to monitor antibody development.

Antibodies confer protection in the overwhelming majority (>90%) of seropositive individuals, with the caveat that the longevity of those antibodies has yet to be determined.^1^ Further, many of the approved SARS-CoV-2 vaccines in use confer high levels of protection against SARS-CoV-2 by provoking a brisk humoral response.^2^ The presence of antibodies has also been the basis for selected therapeutics such as COVID-19 convalescent plasma (CCP)^3^ and monoclonal antibodies against COVID-19.^4^ A determination of serostatus is predictive of response to treatment with these therapies. For example, those who are seronegative at diagnosis have been found to have a significant decrease in hospitalization rate following monoclonal antibody therapy; by contrast, seropositive patients do not benefit significantly from monoclonal antibody therapy.^4^ Similarly, CCP appears to be optimally beneficial when high titer units are provided early in disease course.^5,6^

Recent studies have shown that individuals who have recovered from COVID-19 may only require a single dose of vaccine to confer comparable protection to naïve individuals following receipt of two vaccine doses.^7^ Modification of existing vaccine policy accordingly could reduce the total amount of vaccine doses needed to achieve herd immunity. Nonetheless, it would require a rapid method to screen individuals for antibodies (i.e. to confirm prior infection). To date, the rapid SARS-CoV-2 antibody tests approved under emergency use authorization are lateral flow assays, whose performance characteristics have been highly variable.^8^ Furthermore, lateral flow tests do not offer any quantitative data on antibody levels; the latter are important given the wide range of antibody responses, particularly following mild SARS-CoV-2 infection.^9^

We sought to develop a point-of-care test for SARS-CoV-2 antibody detection based on hemagglutination, leveraging a test platform that is already routinely used at the point-of-care for determination of blood types. Previous work that demonstrated proof of concept for hemagglutination-based SARS-CoV-2 antibody detection, but requiring a one-hour incubation time and pipetting in a 96-well plate.^10,11^ We also sought to determine if the test could yield a semi-quantitative readout of antibody levels among COVID-19 recovered patients within few minutes, which would represent the first rapid SARS-CoV-2 serology test to do so.

## Materials and Methods

### Design and construction of hemagglutination, point-of-care test

Previous studies by our group and a second group have demonstrated the capability of using of fusion protein of an antibody against a red blood cell antigen connected to the receptor binding domain (RBD) of SARS-CoV-2 to detect antibodies against RBD in patient plasma.^10,11^ We adapted dry hemagglutination cards, which are currently used in countries across the world in a room temperature stable kit for rapid, point-of-care testing for ABO and Rh-blood types. We collaborated with Eldon Biologicals (Gentofte, Denmark), who currently sells cards with dried antibodies (EldonCards) to detect ABO and Rh for blood typing. The hemagglutination card kits comes with components of a lancet to elicit blood, a dropper to add water to the platform, as well as stirring sticks to develop the assay, which we utilized in our testing (**Supplementary Figure 1A)**. Each EldonCard test circle has dried antibodies to the target RBC antigen to trigger hemagglutination and typing determination (**Supplementary Figure 1B**). We repurposed these cards for COVID-19 antibody detection by formulating the cards instead with the IH4-RBD fusion protein, previously described.^11^

As outlined in **Figure 1**, we took a fusion protein from one of the studies, IH4-RBD,^11^ and dried it onto a hemagglutination card to formulate the test. The IH4-RBD fusion protein was obtained from Absolute Antibody (Oxford, United Kingdom) as gift of the investigators (Alain Townsend) of the previous study.^11^ 533.2 ng of IH4-RBD protein was dissolved in a proprietary buffer and placed onto the card. The cards were then heated to leave a dried protein mixture on the card, which is stable at room temperature and can be packaged and shipped. Addition of water solubilizes the fusion protein, and addition of blood containing COVID-19 antibodies facilitates cross-linking of RBC’s, which after stirring, can be observed macroscopically (**Figure 1**).

**Figure 1.**
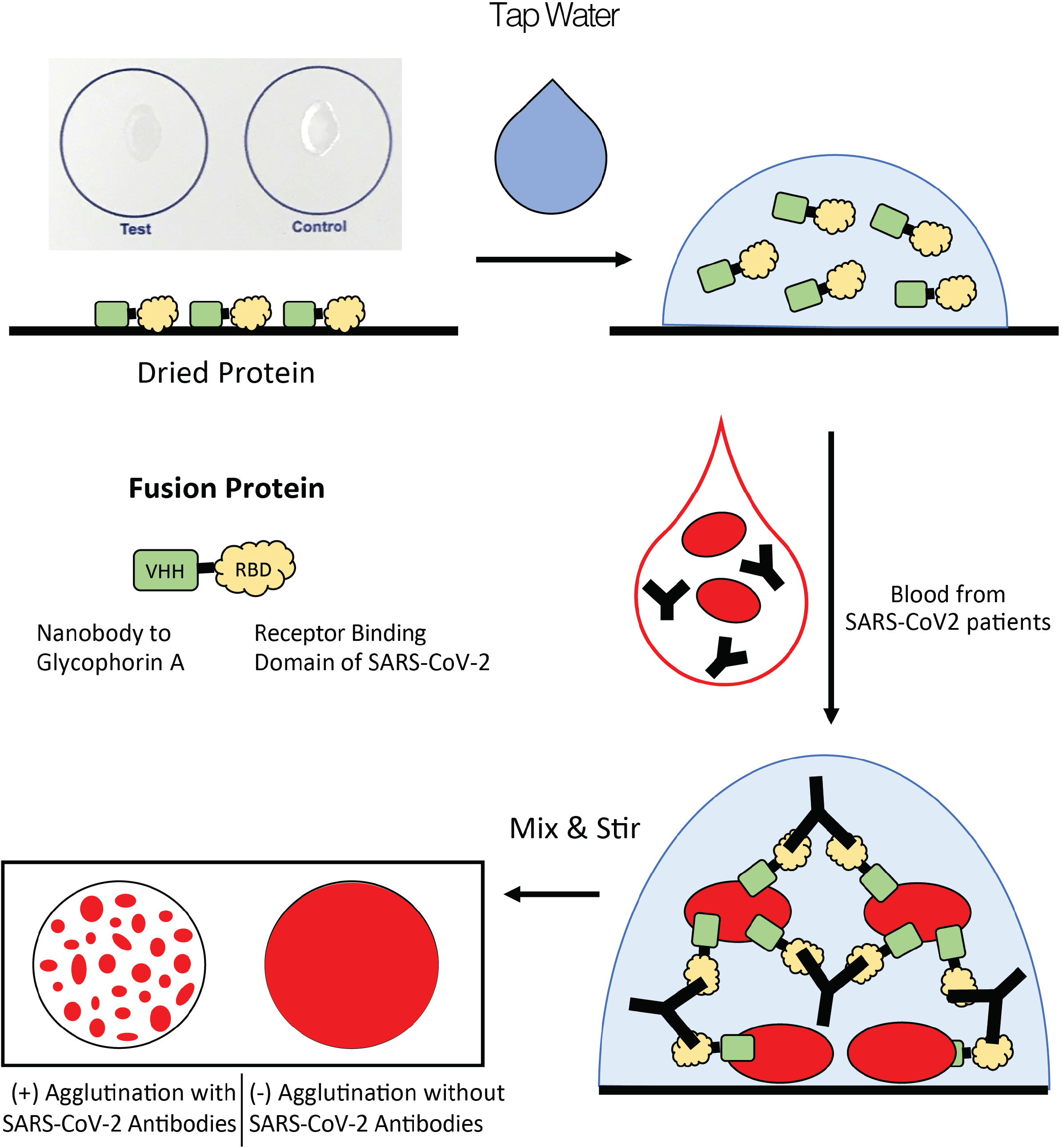
Mechanism of a dry card assay for hemagglutination-based detection of SARS-CoV-2 antibodies. A fusion protein consisting of a nanobody targeting human glycophorin A and the receptor binding domain (RBD) of SARS-CoV-2 was previously described.^11^ The fusion proteins are dried onto a non-water absorbent card, remaining stable at room temperature indefinity in a desiccant pouch. For testing, the fusion proteins are resuspended in a water droplet, followed by the addition of whole blood containing antibodies and RBCs. Stirring facilitates cross-linking of large aggregates of RBCs, which are visible by the naked eye.

### Clinical Specimens tests

The characteristics of the study population and associated clinical specimens have previously been described.^8,12^ In brief, the convalescent SARS-CoV-2 samples had been collected from COVID-19 patients who were confirmed positive by reverse transcription polymerase chain reaction (RT-PCR) at least 28 days prior to specimen collection (mean 45 days, SD 7.5). The pre-pandemic control samples were collected from a prior study of patients who presented to the Johns Hopkins Hospital Emergency Department with symptoms of an acute respiratory tract infection between January 2016 and June of 2019 as part of the Johns Hopkins Center for Influenza Research and Surveillance study. An analysis was conducted of the stored samples and data from those studies. No additional samples were collected for the current study.

### Sample preparation

Type O, Rh-negative packed red blood cells (pRBCs) were obtained from Tennessee Blood Services and provided by Biochemed Services (Winchester, Virginia). On receipt, the pRBCs were stored at 4°C and used entirely within 28 days of collection. All RBCs were washed with phosphate-buffered saline (PBS) to remove any residual plasma. Washed pRBCs were combined with frozen plasma to reconstitute “whole blood” with ∼40% hematocrit after combining pRBCs and frozen plasma.

### Testing protocol

For each test, 20 μL of tap water was placed onto the dried protein spot within the test circles on the card to dissolve the protein. Reconstituted whole blood (20 μL) was then added to the spot. The fluid of water and blood was mixed with a plastic Eldon Stick, spreading the liquid completely within the circle to make sure that the dissolved protein was mixed well with blood on the card. The card was tilted for 10 seconds in each 90 degree direction (4 times in total) and allowed to incubate on a flat surface for 3 minutes. The card was then tilted again as before in each 90 degree direction. Demonstration and validation of the test with a vaccinated individual is shown in **Supplementary Video 1**.

Tests were interpreted according to similar protocols established for scoring hemagglutination in EldonCard blood typing assays. The tests were assigned scores of 4, 3.5, 3, 2.5, 2, 1, and 0. The scores of 1 and 0 were assigned as negative results, where a score of 2 or higher was a positive test result. Tests were interpreted both during tilting of the card, as well as interpretation on a flat horizontal surface, since weak agglutinations could be appreciated in certain cases more easily with the liquid droplet on the side.

### Serological assay testing using commercial assays

CoronaChek SARS-CoV-2 lateral flow assay and the Euroimmun IgG Spike ELISA were performed in accordance with the manufacturers’ instructions and tests results previously reported on a different study on the same specimens.^8,12,13^ Neutralizing antibody assay (titer and AUC determination) data were made available; their acquisition has previously been described.^12,14^

### Statistical Analyses

Sensitivity of the assays was calculated among COVID-19 convalescent individuals and specificity was calculated among pre-pandemic population. 95% Clopper-Pearson Confidence Intervals (CI) of sensitivity and specificity were also computed. Statistical comparisons made between agglutination scores and ELISA optical density and neutralizing antibody titers or AUC were made using non-parametric Mann-Whitney tests. Statistical significance was set at P<0.05. All statistical analyses were performed using GraphPad Prism 9.0.0 for Mac, GraphPad Software, San Diego, California USA, www.graphpad.com.

### Human subjects

The parent studies of the collected patient samples were approved by The Johns Hopkins University School of Medicine Institutional Review Board (IRB00247886, IRB00250798, and IRB00091667). All samples were deidentified prior to testing in the current manuscript, and the original studies were conducted according to the ethical standards of the Helsinki Declaration of the World Medical Association.

## Results

### Hemagglutination test performance against clinical samples

Plasma samples obtained from COVID-19 convalescent (n=200) and pre-pandemic individuals (n=200) were reconstituted with O-negative blood to a hematocrit ∼40%, approximating whole blood for the hemagglutination test. The testing protocol matched the protocol that is currently used in ABO typing with the EldonCard resulting within 5 minutes. The sensitivity for detecting antibodies was 87.0% (CI: 81.5% - 91.3%). By comparison, the FDA-approved Euroimmun Spike IgG ELISA test showed a sensitivity of 86.5% (CI: 81.0% - 90.9%), while a high-performing RBD-based lateral flow assay, CoronaChek,^15^ had a sensitivity of 84.5% (CI: 78.7% - 89.2%) on the same 200 samples (**Table 1**). Specificity for the hemagglutination test was calculated at 95.5% (CI: 91.6% - 97.9%), which was not statistically lower than the Euroimmun ELISA, 97.3% (CI: 95.5% - 98.5%), and CoronaChek, 99.0% (97.8% - 99.6%), respectively. An additional cohort of 16 lateral flow assays was also compared in sensitivity to the hemagglutination test on a smaller set of samples, with the hemagglutination test showing similar sensitivity (data not shown).^8^

**Table 1.** Hemagglutination-based SARS-CoV-2 antibody assay performance Sensitivity and specificity are presented for the hemagglutination test using 200 samples of PCR-confirmed COVID-19 patients and 200 pre-pandemic samples of patients with acute respiratory symptoms. Results of a regulatory-approved Euroimmun Spike IgG ELISA test and RBD-based CoronaChek lateral flow test on the same samples are also presented for comparison. Specificity results are presented for an equivalent bank of pre-pandemic samples, although not all samples overlap between the three groups. Borderline samples on ELISA were called positive, and faint samples on lateral flow assay were called positive. Confidence intervals are presented within parentheses.

| Antibody test | Sensitivity (CI) | Samples | Specificity (CI) | Samples |
| --- | --- | --- | --- | --- |
| Hemagglutination Test | 87.0% (81.5% - 91.3%) | 174/200 | 95.5% (91.6% - 97.9%) | 191/200 |
| Euroimmun Spike IgG ELISA | 86.5% (81.0% - 90.9%) | 173/200 | 97.3% (95.5% - 98.5%) | 502/516 |
| CoronaCheck Lateral Flow Assay | 84.5% (78.7% - 89.2%) | 169/200 | 99.0% (97.8% - 99.6%) | 574/580 |

We next scored all agglutinations observed on the card across 200 samples tested, building off of the previous scoring system of hemagglutination developed by Eldon for ABO typing. As shown in **Figure 2**, the highest agglutination was scored at 4, wherein large clumps of RBCs are seen with few residual free cells, to 0 wherein no reaction is observed. The agglutination scores of 0 and 1 were termed to be negative, while any score at 2 or above was positive. Scoring is presented as the cards resting on a horizontal surface in **Figure 2**, as well as slanted after final mixing (**Supplementary Figure 2**). In the latter scenario, it can often be easier to see small agglutinations without the large liquid droplet of un-agglutinated RBCs obscuring the view, as well as the kinetic movement of these agglutinations across the test circle. In an agglutination score 1 field, there can be some small number of agglutinations observed, but these are usually very few and often fixed to card, and do not move like most agglutinations in a 2 score.

**Figure 2.**
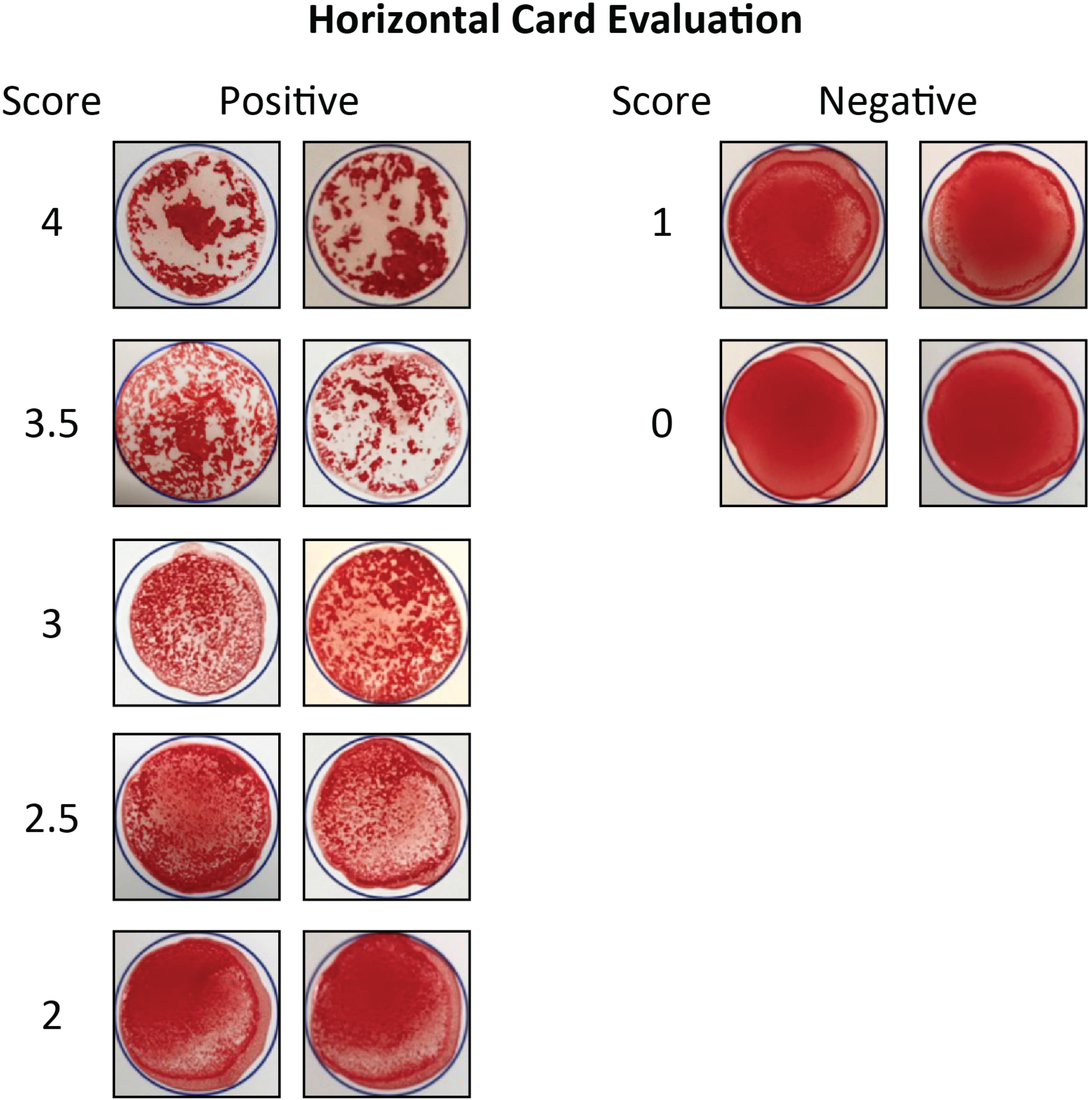
Hemagglutination can be scored for reaction strength. Scores were developed to quantify the degree of agglutination observed across COVID-19 convalescent samples. Cards are depicted horizontal on table surface after testing. Strong agglutinations (ex: 4) quantified the majority of red blood cells sticking together, with a white background without unbound cells. Weaker reactions (ex: 2) had smaller, but frequent agglutinations. The scores of 0 and 1 were deemed negative for the purpose of the test.

Across the 200 recovered COVID-19 patients, a range of different agglutination scores were observed (**Figure 3A**). Interestingly, relatively few patients achieved the highest levels of agglutination 4 and 3.5, while 47% patients had borderline studies (2-2.5). We sought to confirm that these differences in agglutination were related to the antibody concentration in the plasma, which would influence the amount of RBC cross-linking observed. We took a patient sample that scored a 4 agglutination and performed a serial dilution in order to assess if agglutination decreased. A decline in agglutination correlated progressively with more dilute samples (**Figure 3B**), with a 1:10 dilution scoring a clear, but weak reaction, while a 1:50 dilution did not show a clear reaction after 5 minutes. Thus, the agglutination results correlate with antibody concentration.

**Figure 3.**
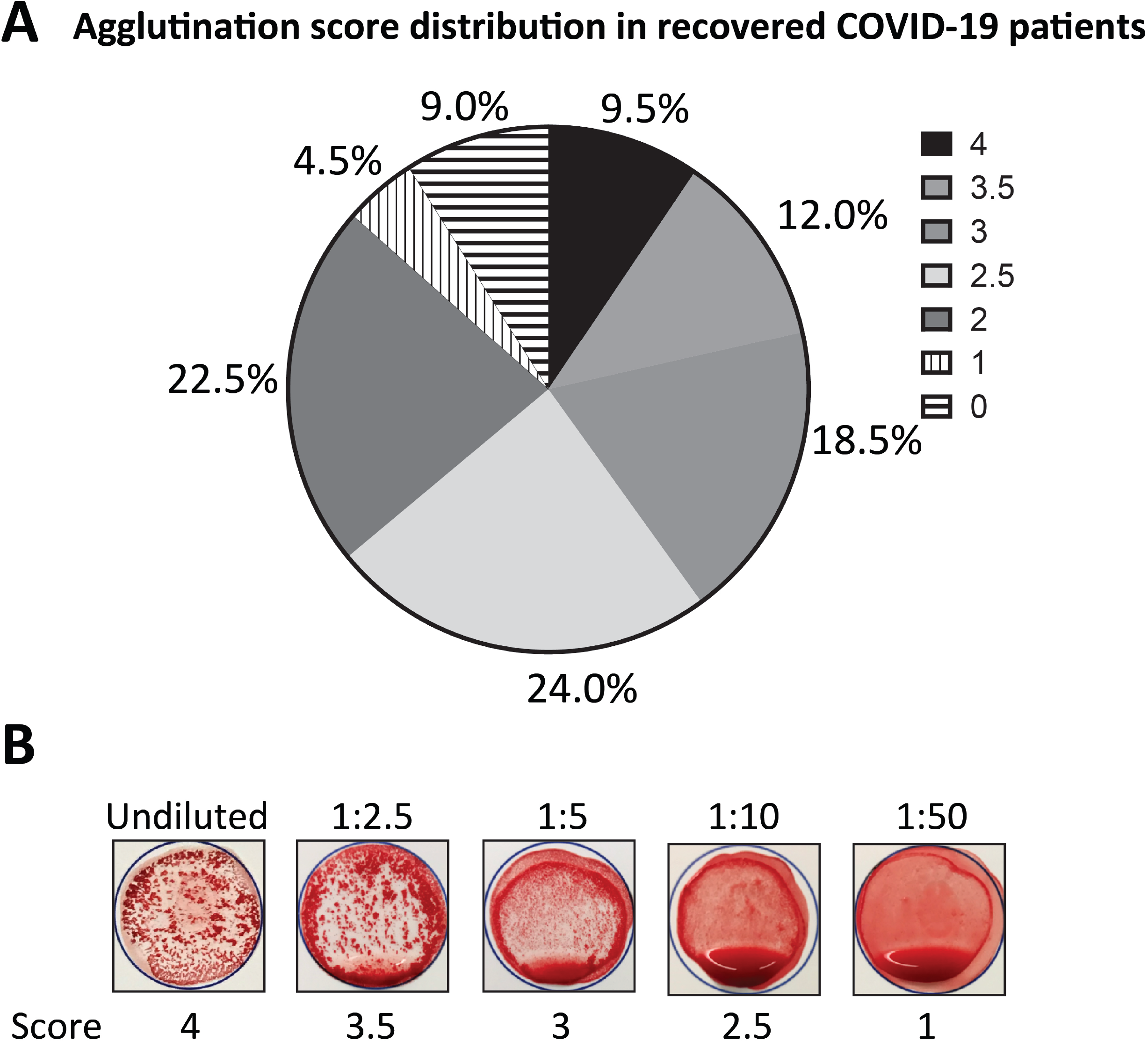
COVID-19 recovered patients exhibit a distribution of agglutination scores that correlate with the dilution of COVID-19 convalescent plasma. (A) Agglutination scores for 200 patient samples were tabulated, and percentages for each agglutination score provided. Agglutination scores of 1 and 0 were deemed negative and are stripped for distinction from the positive test results in solid color. (B) A sample with a strong agglutination (4) was obtained, and the plasma progressively diluted with pre-pandemic plasma. The same amount of RBC’s was added to all conditions. Agglutinations are depicted in the titled position, and were clearly seen down to 1:10, while 1:50 only had very weak agglutinations below the assay cutoff.

The relationship between agglutination score and traditional ELISA and neutralizing antibody assays against SARS-CoV-2 was next determined. There was a correlation between the optical density (OD) of the Spike IgG ELISA assay and agglutination score, despite the hemagglutination assay only containing RBD, which is a small portion of the Spike protein (**Figure 4A**). The RBD is a major target of neutralizing antibodies, so we examined agglutination score versus neutralizing antibody levels. There was a general correlation with increasing agglutination score and higher neutralizing antibody levels for both the AUC (**Figure 4B**) and endpoint dilution titer (**Figure 4C**) against the virus. Notably, agglutination scores of 1 and 2 had no difference in neutralizing antibodies, while strong agglutination scores 3 or higher were clearly defined by higher neutralizing antibody levels.

**Figure 4.**
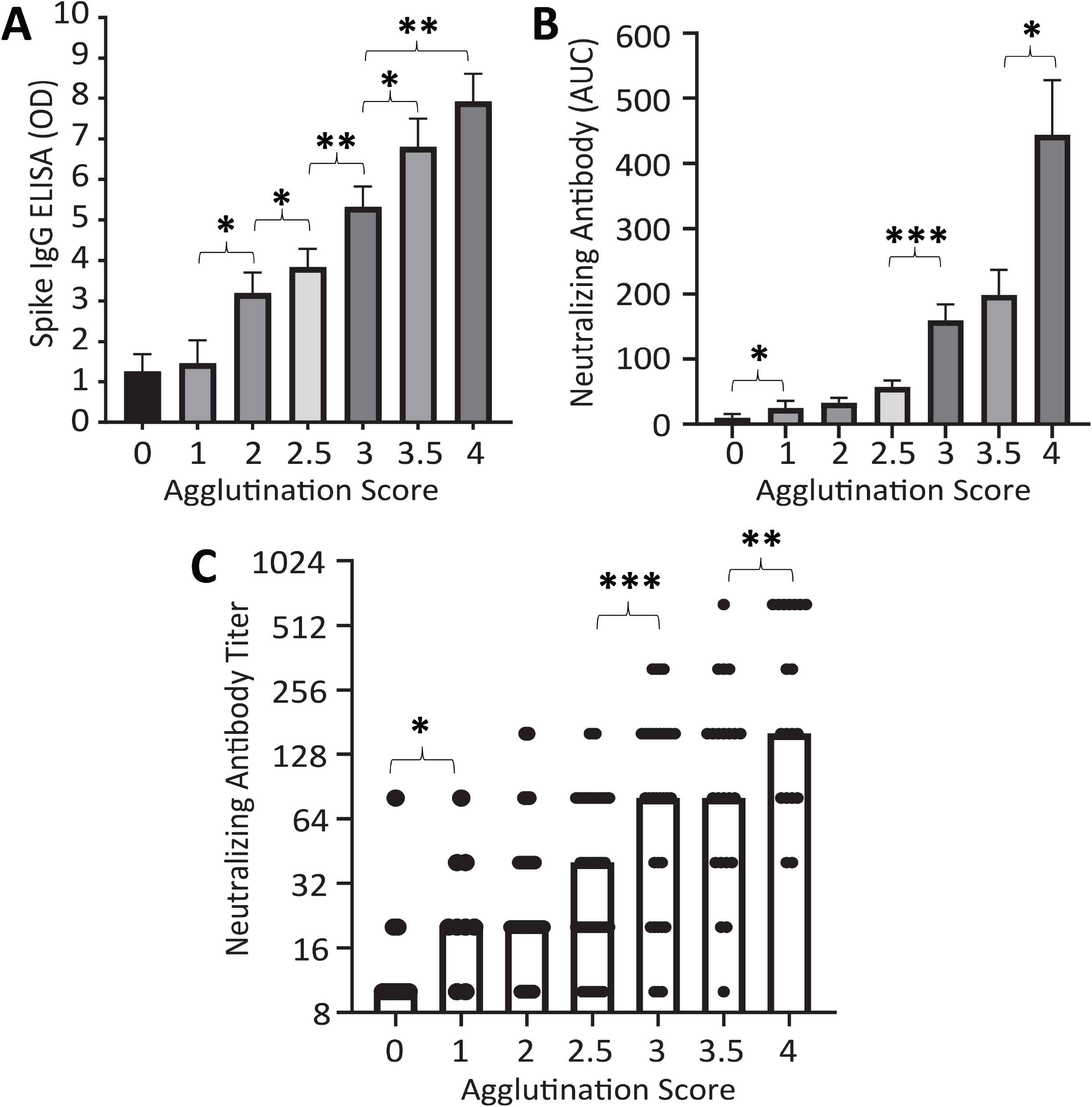
Agglutination scores correlate with ELISA and neutralizing antibody titer assays. (A) The optical density (OD) of the Euroimmun Spike IgG ELISA (1:101 dilution) was categorized for each agglutination score. (B) Neutralizing antibody AUC (area under curve) was quantified for each specimen and plotted against the respective agglutination score. (C) A plot of neutralizing antibody titers at different agglutination score is presented, wherein each dot represents a single sample, and each bar represents the median among the samples. Statistical analysis was performed using non-parametric, one-tailed Mann-Whitney U tests. Statistical significance (* p<0.05, ** p<0.01 *** p<0.001)

### Analysis of false positive and negative samples

We next sought to determine how the assay time influenced the observed test performance, given our experience that many strong agglutination reactions were observed after just the first series of card tilting and blood mixing. In a smaller cohort of 73 tests within the 200 samples, we scored the test after initial tilting as well as after the complete assay time. We found that the sensitivity of the assay decreased to 60.3% if only one-minute of assay time was leveraged (**Figure 5A**). The additional incubation time led to another 23.3% of samples becoming positive, although the agglutination scores of these tests were almost all 2 (94.1%) (**Figure 5B**). Interestingly, the neutralizing antibody titers of these weak reactions requiring additional incubation time were all very low and were significantly lower than the neutralizing antibody titers for similarly weak reactions (2-2.5) that were initially visible after 1-minute (**Figure 5C**). Together, this indicates increased sensitivity, but lower functional utility in increasing assay time.

**Figure 5.**
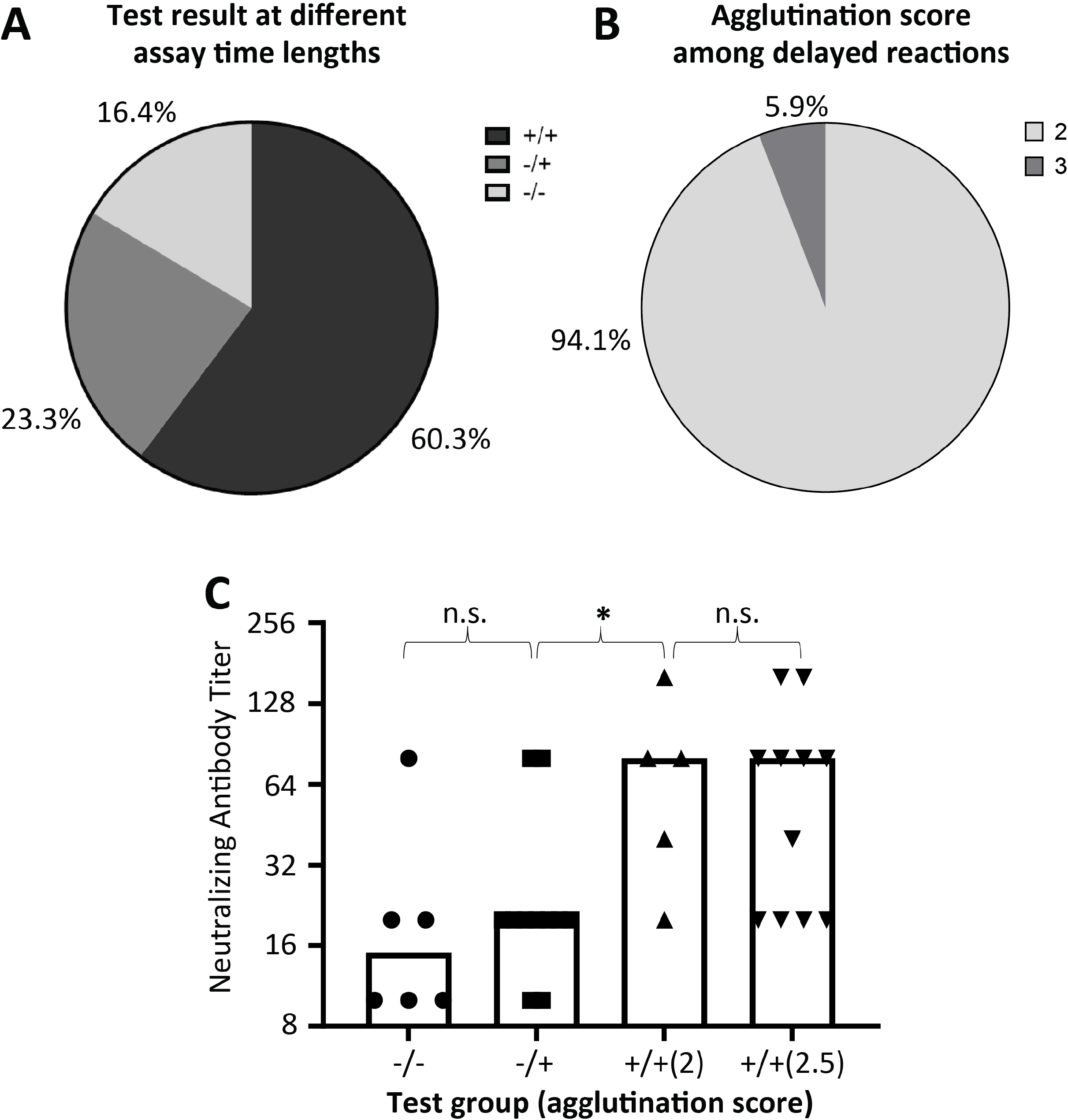
Specimens requiring longer assay time to yield agglutination have low agglutination scores and neutralizing antibody titer. A subset of convalescent COVID-19 samples (n=73) was scored after the first round of card tilting, followed by three minutes incubation and a second round of card tilting for a second score. (A) The distribution of samples is presented that were positive already at the first round (+/+), positive after the second round (-/+), and negative after the complete assay (-/-). (B) The agglutination scores of -/+ samples are provided, demonstrating almost all scored a 2 agglutination score. (C) The neutralizing antibody titers are presented for negative (-/-), delayed agglutination (-/+), and fast (+/+) samples with low agglutination scores, 2 and 2.5, respectively. The latter would correspond to the ultimate agglutination scores observed by -/+ samples. Each point represents a single sample, and each bar represents the median of the group. Statistical analysis was performed using a non-parametric, Mann-Whitney U test. Statistical significance (* p<0.05) n.s. = non=significant

The false positive samples in the pre-pandemic samples were next interrogated. Among the 9 false positives, 6 required the additional incubation time to become positive (**Figure 6A**). The agglutination scores among the false positives were also weak, with only 2 out of 9 registering a score of 3 (**Figure 6B**). If the assays were limited to just interpretations after 1-minute, the specificity would increase to 98.5% (197/200). The false negatives among the ELISA, hemagglutination, and CoronaChek assays were next compared. While there were no statistical differences between tests, the lateral flow test had a trend toward higher levels of neutralizing antibodies completely missed (**Figure 6C**). Interestingly, one sample measured 1:320 neutralizing antibody titer, a score of 4 by agglutination, but was negative on the CoronaChek test.

**Figure 6.**
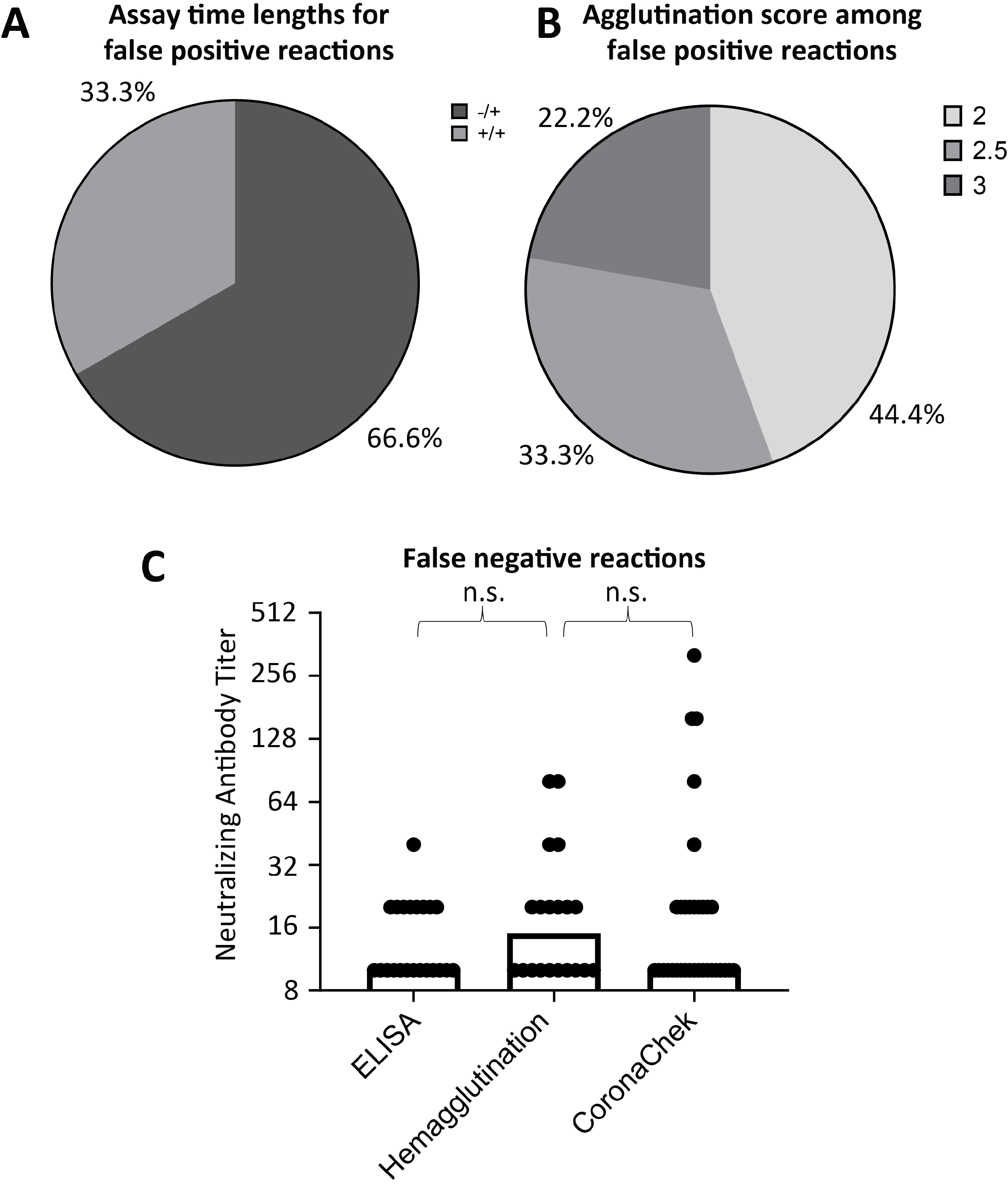
False positive samples on hemagglutination test yield weak agglutinations, while high-titer false negative agglutinations are rare. 9 samples that were false positives among the pre-pandemic, emergency department samples were further analyzed. (A) The distribution of samples is presented that were positive already at the first round (+/+) versus positive after the second round (-/+). (B) The distribution of agglutination scores among false positive samples are also demonstrated, generally showing very weak agglutination. (C) Neutralizing antibody titers are presented for false negative samples among the hemagglutination-based test, Euroimmun Spike IgG ELISA, and the CoronaChek lateral flow assay. Each point represents a single sample, and each bar represents the median of the group. Statistical analysis was performed using a non-parametric, Mann-Whitney U test. n.s. = non=significant

## Discussion

In this study, we established a new point-of-care SARS-CoV-2 antibody testing platform that distinguishes itself from the commonly used lateral flow assays given its being semi-quantitative for detecting antibodies. We found that this hemagglutination-based assay has a more rapid turnaround time than lateral flow assays (5 minutes vs 15 minutes respectively) and ELISA assays (24-48 hour turnaround time) by comparison and can achieve similar sensitivities. Moreover, a technically equivalent test is sold commercially by Eldon Biologicals for blood typing, which should aid in the manufacturing and regulatory approval.

Our results found 87.0% sensitivity among COVID-19 recovered patients, which compared well to leading ELISA (86.5%) and lateral flow assays (84.5%), which are already in use. Our sensitivity was similar to the prior study using the IH4-RBD fusion in a 96-well plate with one-hour incubation, which found a 90% sensitivity.^11^ The slight differences in sensitivity between the two assays may result from the longer incubation time and gravity in a 96-well, U-bottom plate facilitating RBC aggregation. While many ELISA and lateral flow assays use the whole spike protein, we chose to use the spike RBD for this test, since it is the main target of neutralizing antibodies, which should provide protective immunity. RBD has been employed as the target antigen in ELISA^16^ and lateral flow tests,^8^ respectively. Some results from RBD-based ELISA tests have found high sensitivity (98%) and specificity (100%)^17^ ; sensitivity of 96% and specificity of 99.3%^18^ 88% sensitivity and 98% specificity.^19^ The original CoronaChek RBD-based lateral flow assay description reported a sensitivity of 88.66% and specificity of 90.63%, differing from markedly from the values in this publication.^15^ The differences between all these studies are likely driven by variable patient symptoms and emphasizes the need for comparison of tests on the same set of samples.

A key advantage of our test is the semi-quantitative readout of antibody levels, which is unique among all point-of-care COVID-19 serology assays on the market today. We observed a correlation between agglutination score and neutralizing antibody titer, which has also been seen in RBD-based ELISA assays where a correlation with neutralizing antibody titer was found.^17^ The ability to interpret an RBC agglutination pattern for semi-quantitative determination was previously used in the SimpliRED test, evaluating D-dimer levels at the point-of-care.^20^ Importantly, the correlation between neutralizing antibody levels and agglutination can also help refine use cases for the test in the future, such that scores lower than 3 are determined to lack substantial immunity. Of note, preliminary testing of vaccinated individuals with our hemagglutination test has shown agglutinations of 3.5 or 4, matching the uniformly high levels observed in clinical trials

The wide distribution of agglutination scores we observed matches the variability in antibody responses among COVID-19 patients wherein not every patient recovered from COVID-19 produces antibodies at high levels or in some cases at all, particularly among non-hospitalized cases with mild to no symptom,^21^ while hospitalized patients are noted to have significantly higher antibody and neutralization titers.^22^ the testing performance is reported is strongly dictated by antibody levels in the specimens, which is in turn dictated by the patient population, with severe disease patients having higher antibody levels than mild cases.^23^

This study had limitations. The specificity in our assay (95.5%) was lower than the 99% reported using the same fusion protein previously, and also lower than the aforementioned RBD-based ELISA tests (specificity of 100%^17^ and 99.3%^18^). The reason for lower specificity is uncertain but is likely multi-factorial. The manufacturing of a dried protein on the card may yield fusion protein clumping not seen in protein in solution in the prior report.^11^ Another consideration is that the prior study^11^ tested healthy donors as a control, while all negative control samples in our assay were patients with acute respiratory illness, including a subset with active seasonal coronavirus infection. While the sequence identity is only ∼20% shared between the viruses^24^ it’s possible that even weakly cross-reactive antibodies could achieve binding at high concentrations. Cross-reactivity has been suggested as a reason for significantly worse specificity of SARS-CoV-2 antibody ELISA assays (90-94% against spike protein)^25^ and CoronaChek lateral flow (96.5%)^13^ in African populations. An important distinction is that ELISA cutoff values for optical density can be optimized for maximal specificity,^26^ whereas the hemagglutination test relies on visual interpretation with more limited nuance. Soluble ACE2 and white blood cells expressing ACE2 may also contribute to false positives via binding to RBD fusion protein. Of note, specificity could be increased up to 98.5% if assay time was reduced to the initial 1-minute of tilting, suggesting that these false positives could be eliminated with further modification of the assay. Moreover, non-reacting tests at 1-minute did not have high levels of neutralizing antibodies, meaning they may have decreased protection anyways.

Among other limitations, our hemagglutination test on a dry card is also limited by the inability to distinguish between IgG/IgM/IgA, though it may be useful to take into all antibody isotypes when assessing total neutralizing antibody levels. A further limitation of our approach is that it requires visual interpretation by the user, which may be variable. To solve this, automated visual algorithms leveraging cameras on phones could be used. One study has explored the potential for interpreting strong and weak lines from lateral flow assays to correlate with antibody levels,^27^ but such strategy hasn’t been translated into commercial instructions and is visually more subtle than the agglutination results presented in this study.

In summary, we have developed a new platform for SARS-CoV-2 antibody detection, that is faster than current point-of-care devices and offers semi-quantitative information. The simplicity and low cost of the assay could enable its widespread use and a range of applications that include testing in low resource settings, determination of serostatus prior to vaccination, post-vaccination surveillance and travel screening.

## Data Availability

All data is available in the manuscript upon request to the authors.

## Acknowledgements

We would like to thank the Peter Stounbjerg and Kim Kristensen for production of EldonCards and executing pilot testing, and thank Kasper Juul Hedegaard, Hans-Ole Hedegaard, and Michael L. Hansen of Eldon Biologicals for their advice and coordination during the project. We would also like to thank Heather Smetana and Melissa Neally for providing reagents and technical advice. This work was funded by the Johns Hopkins Department of Pathology Fred and Janet Sanfilippo Research Award (R.L.K), and partially supported by the National Institutes of Health, National Institute of Diabetes and Digestive and Kidney Disease [grant R01DK106109] (Z.Z.W).

## Declarations

R.L.K, Y.H., and Z.Z.W. are co-inventors on a patent application related to the fusion protein detection agent for SARS-CoV-2. The authors recruited Eldon Biologicals for the collaboration, but the data and conclusions here are fully the authors and independent from Eldon Biologicals. Beyond testing cards, no formal sponsored research funding was received from Eldon Biologicals.

## Supplementary Data Legends

**Supplementary Video 1. Demonstration of point-of-care testing for SARS-CoV-2 antibody detection with a hemagglutination test**

A vaccinated individual demonstrates the protocol to detect antibodies on the EldonCard. The assay is stopped after the first round of card tilting since a strong agglutination is already observed. Weaker agglutinations would continue with three minutes of incubation time, followed by a second round of card tilting.

Video Link: https://www.youtube.com/watch?v=Uu4oawH4Hho

**Supplementary Figure 1. The EldonCard as a test of point-of-care, hemagglutination for ABO typing**.

The EldonCard (A) is a commercially sold point-of-care test for blood typing. The kit (B) includes an alcohol pad to sterilize a finger tip, and lancet to yield drops of blood. Plastic sticks are included to help collect and stir blood drops. The card is kept in a desiccating pouch for shipping and is stable at room temperature. (C) The Eldon Card itself has antibodies against the ABO and Rh blood groups dried onto spots on the card. Results from testing an A-positive person is shown, demonstrating strong agglutination.

**Supplementary Figure 2. Tilted cards can facilitate agglutination scoring**.

Tilted cards were evaluated for agglutination after the last step stirring step. Strong agglutinations (ex: 4) quantified the majority of red blood cells sticking together, with a white background without unbound cells. Weaker reactions (ex: 2) had smaller, but frequent agglutinations, that were observed during card tilting. The scores of 0 and 1 were deemed negative for the purpose of the test.

