## Supplementary figures and images for "A hemagglutination-based, semi-quantitative test for point-of-care determination of SARS-CoV-2 antibody levels"

### Supplemental Figures

**Supplemental Figure 1**

**A**

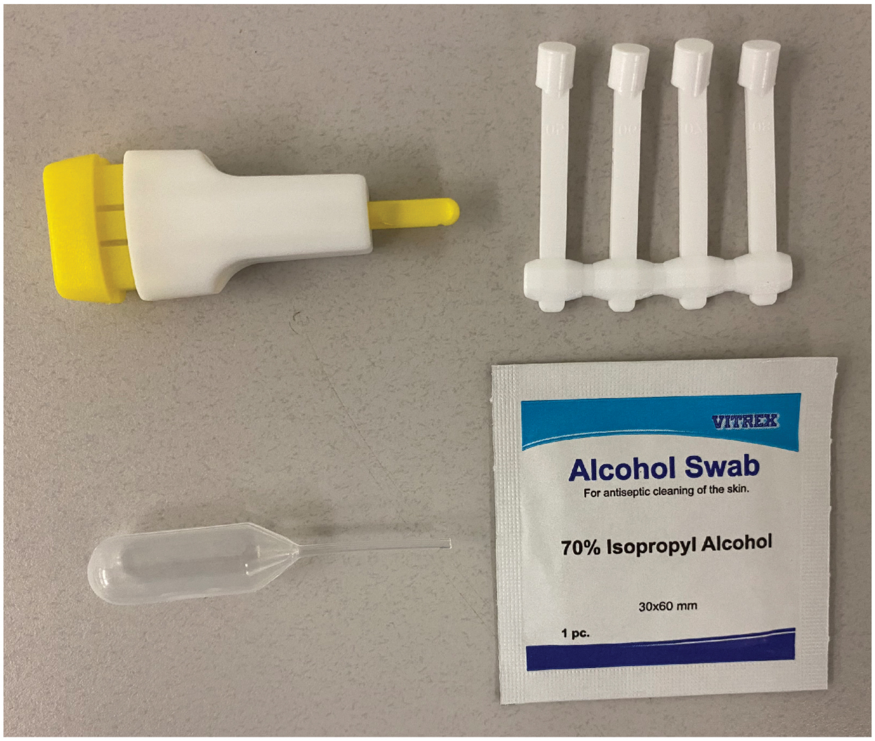


**B**

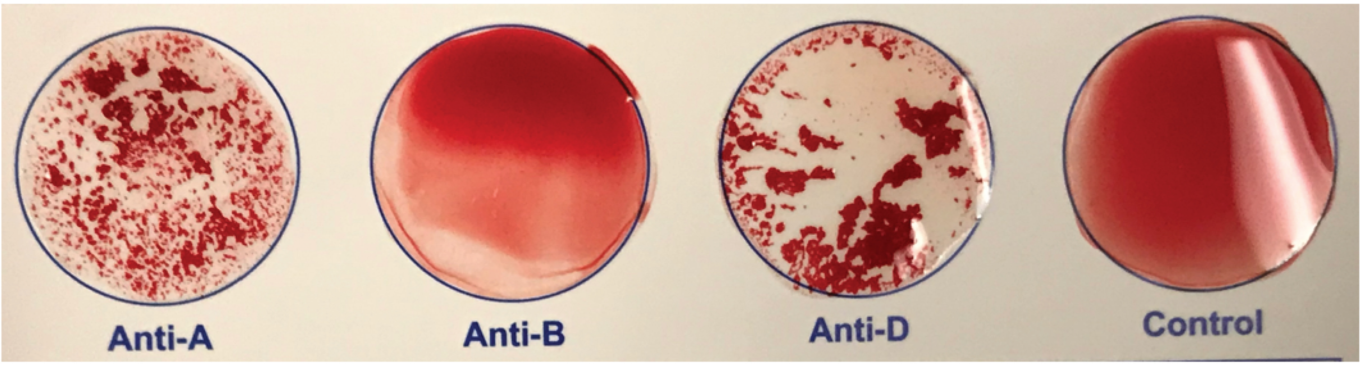

#### Supplemental Figure 2

##### Tilted Card Evaluation

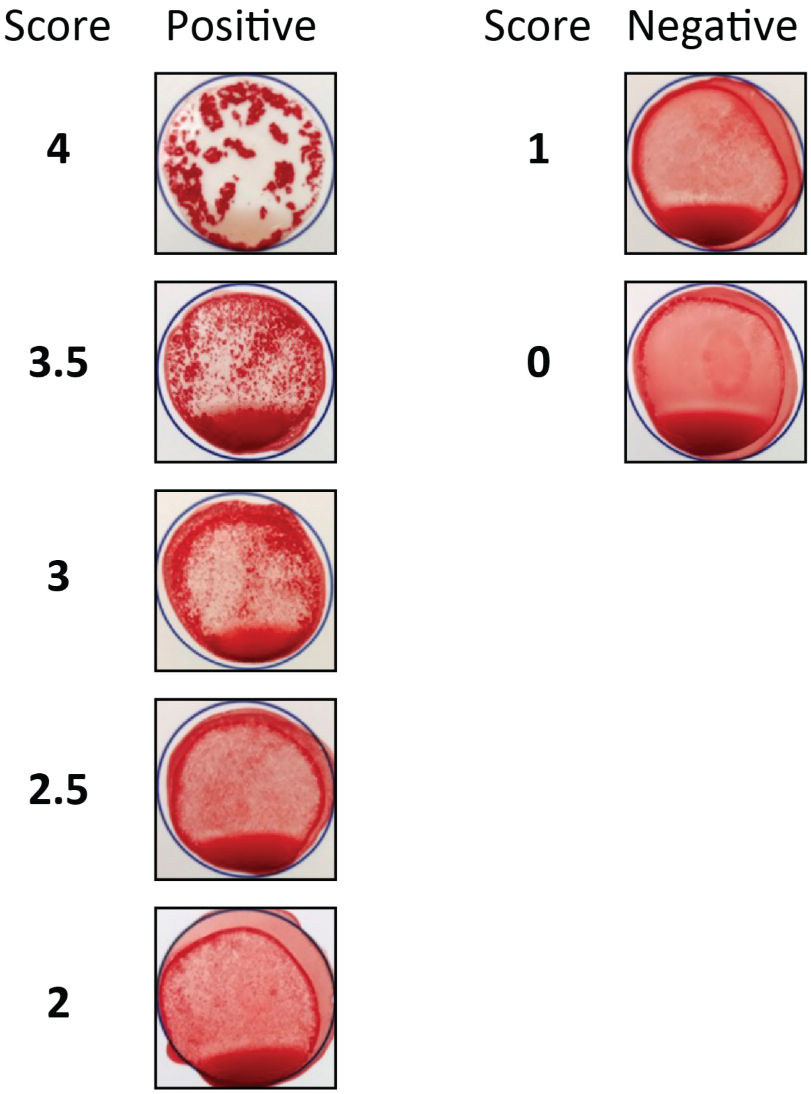
